# Investigation of autistic traits in university students and the intervention effects of Tai Chi practice: a non-randomized controlled trial

**DOI:** 10.1101/2025.09.23.25336500

**Authors:** Yuan Zhao, Yue Wang, Weijiao Zhong, Qiqi Xing

## Abstract

Background: Autistic traits in the general population are linked to emotional regulation difficulties and aggression, affecting psychological adaptation. Tai Chi, a mind–body exercise, may improve psychological well-being. Methods: This study adopted a mixed-methods approach combining questionnaire surveys and experimental intervention. In Study 1, the Autism Spectrum Quotient (AQ) was used to assess the current status of autistic traits among university students. In Study 2, a group of university students with high levels of autistic traits participated in an 8-week Tai Chi intervention. Changes in autistic traits, emotional regulation self-efficacy (ERSE), and aggressive behavior (Reactive-Proactive Aggression Questionnaire, RPQ) were measured before and after the intervention. Results: The results indicated that overall autistic traits among university students were at a moderate level. Males scored significantly higher than females on the AQ total score and on the dimensions of communication and imagination. Following the Tai Chi intervention, participants showed a significant reduction in overall AQ scores, particularly in attention switching and attention to detail. Conclusion: Tai Chi training demonstrates positive effects on emotional regulation and behavioral control among university students with autistic traits. Future research should further explore the underlying mechanisms and broader applicability of such interventions.

## Introduction

Autistic traits refer to behavioral characteristics such as social withdrawal, direct speech, lack of imagination, preference for solitude, and difficulty in emotional expression [1]. Clinically, these traits are often accompanied by restricted interests, stereotyped and repetitive behaviors, as well as other potential psychological issues. They can be assessed using the self-report Autism-spectrum Quotient (AQ) (Autism-spectrum Quotient, AQ) [2]. Research has shown that autistic traits are widely distributed in the general population and exist along a continuum. As a quantifiable personality dimension, these traits may develop into autism spectrum disorder (ASD) when expressed at an extreme level [3]. There is a significant gender disparity in the prevalence of ASD, with a male-to-female ratio of approximately 3–4:1 [4]. According to the 2022 Report on the Development of Autism Education and Rehabilitation in China (IV), the number of individuals with autism in China has exceeded 10 million, with an annual increase of nearly 300,000 new cases. These figures indicate that the prevalence of autism is rapidly rising worldwide, making it a growing global public health concern [5]. In addition to the core clinical symptoms, individuals with autism often exhibit comorbid conditions and problematic behaviors. Among these, aggressive behavior not only severely affects daily functioning but also hinders long-term development and increases the risk of symptom deterioration [6].

In recent years, non-pharmacological psychological interventions have garnered increasing attention for their role in enhancing individual mental health. Tai Chi, a traditional Chinese mind-body exercise rooted in classical culture, has become increasingly favored by psychological intervention researchers and practitioners. Based on the philosophical principles of yin-yang, Tai Chi emphasizes the integration of movement and stillness, unity of body and mind, intentional guidance, and breath regulation. It not only carries profound cultural and philosophical significance but also demonstrates notable physiological and psychological therapeutic effects [7]. Long-term practice of Tai Chi has been shown to effectively alleviate anxiety and depressive symptoms, enhance subjective well-being and self-efficacy, and thereby help individuals build more positive psychological resources [8]. The practice’s slow pace, fluid movements, controlled breathing, and sustained attentional focus enable individuals to redirect their attention from external stressors to internal experiences, enhance emotional awareness, and promote emotional self-regulation [9]. This integration of physical and mental coordination gives Tai Chi a unique advantage in addressing emotional difficulties, managing impulsive behavior, and relieving psychological stress. Therefore, Tai Chi is not merely a traditional health-preserving technique but also a culturally meaningful and psychologically beneficial intervention method, particularly suited for individuals with limited emotional regulation capacity and poor behavioral control [10]. For college students with autistic traits—a population often facing greater challenges in emotional regulation and social adaptation, as well as a higher risk of aggressive behavior with limited means of coping—this study attempts to introduce Tai Chi as a psychological intervention. The goal is to explore its potential in enhancing emotional regulation self-efficacy and reducing aggressive behaviors. On one hand, this approach may offer a sustainable path for psychological adjustment in this population, alleviating their adaptation difficulties within the university context; on the other hand, it contributes to the development of a culturally grounded psychological intervention model, expands the application of traditional exercises to groups with special personality traits, and enhances the diversity and scientific foundation of mental health education in higher education institutions.

Among individuals with autistic traits, aggressive behavior is both common and complex in its manifestations. Aggression is defined as any behavior that intentionally inflicts harm or causes pain to others, often against their will, with intensity, frequency, severity, or duration that exceeds what is socially tolerable [11]. Based on different antecedents and functions, aggression can be classified into proactive aggression and reactive aggression [12]. Proactive aggression refers to deliberate, goal-directed, and instrumental actions intended to achieve specific outcomes. Individuals who engage in proactive aggression often experience positive feelings when their goals are accomplished, and such behaviors are frequently associated with criminal conduct and deficits in moral reasoning. In contrast, reactive aggression is a defensive or retaliatory response to perceived provocation or threat and is typically accompanied by intense emotional states such as anger and hostility [13]. Previous studies have shown that aggressive behavior in children with ASD includes both physical forms of proactive aggression and emotionally driven reactive aggression, such as outbursts of anger and hostile retaliation [14]. Moreover, the presence and intensity of aggressive behaviors have been found to correlate with the severity of core autism symptoms and, in turn, may exacerbate stereotyped and repetitive behaviors [2].

These aggressive behaviors are often closely associated with deficiencies in emotional regulation abilities. Emotional regulation self-efficacy refers to an individual’s perceived ability and belief in effectively managing and regulating negative emotions such as anger, anxiety, and frustration. As a critical psychological resource, it not only influences the process of emotion control itself but also significantly affects how individuals respond to external stressors and whether they are prone to exhibit aggressive behaviors [15]. Research has shown that individuals with low emotional regulation self-efficacy are more likely to lose control under heightened emotional arousal or situational stress, thereby demonstrating more frequent and intense aggression. Conversely, those with high emotional regulation self-efficacy tend to regulate impulses more effectively, adjust their emotional responses during adversity, and adopt more rational and socially adaptive behaviors [16]. For individuals with autistic traits, difficulties in recognizing, understanding, and regulating emotions are particularly pronounced. These individuals are especially susceptible to emotional overload when confronted with unexpected or complex social situations [17]. They often lack adaptive emotion regulation strategies and instead exhibit dysfunctional coping patterns, such as avoidance, emotional suppression, or explosive outbursts. Such tendencies not only impair their social interaction skills but also substantially increase the risk of aggressive behavior [18].

In summary, given the psychological adjustment challenges faced by university students with autistic traits—particularly the association between frequent aggressive behaviors and inadequate emotional regulation—developing scientifically grounded, effective, and sustainable intervention strategies holds significant practical importance. Accordingly, this study was conducted in two phases: the first phase investigated the current status of autistic traits among university students; the second phase involved the design and implementation of an 8-week Tai Chi intervention to examine its effects on aggressive behavior and emotional regulation self-efficacy in students with high levels of autistic traits. Specifically, the following hypotheses were proposed for the second phase of the study: H1: The 8-week Tai Chi intervention will significantly reduce aggressive behaviors in university students with high levels of autistic traits. H2: The 8-week Tai Chi intervention will significantly improve regulatory emotional self-efficacy in university students with high levels of autistic traits. The overarching aim was to explore the psychological adjustment mechanisms and applied value of traditional exercise-based interventions for populations with distinct personality characteristics.

### Study 1: Investigation on the current situation of autistic traits among college students Participant

A convenience sampling method was employed to distribute questionnaires online to university students across China. Participants were drawn from 23 provinces, municipalities, and autonomous regions, including Shandong, Guangdong, Sichuan, and Heilongjiang. The participant recruitment period for this study was from September 1, 2024 to September 20, 2024. A total of 973 responses were collected. After excluding invalid questionnaires—such as those completed carelessly or with excessively short response times—906 valid responses were retained (549 male, 357 female), yielding a validity rate of 93.11%. According to a power analysis conducted using G*Power 3.0, assuming a medium effect size (*f²* = 0.15, *α* = 0.05, 1 – *β* = 0.90), the minimum required sample size was 114 participants, indicating that the current sample size was adequate for statistical analyses. All procedures adhered to the ethical guidelines outlined in the Declaration of Helsinki and were approved by the Ethics Committee of Shandong University of Political Science and Law. Participation was entirely voluntary, and informed consent was obtained from all participants prior to the survey and intervention phases. Data collection was conducted under principles of confidentiality and voluntariness. Participants were informed of their right to withdraw from the study at any time without penalty, especially in the event of discomfort or other concerns.

### Research Materials

Participants’ autism traits were assessed using the Autism-Spectrum Quotient (AQ), originally developed by Baron-Cohen et al. (2001) and revised for use in Chinese populations by Zhang et al. (2016). The Chinese version of the AQ has demonstrated good reliability and validity in both clinical and non-clinical samples within China. The full scale comprises 50 items distributed across five sub-scales: social skills (e.g., “I find social situations easy”), attention switching (e.g., “I frequently get so strongly absorbed in one thing that I lose sight of other things”), attention to detail (e.g., “I usually notice car number plates or similar strings of information”), communication (e.g., “I enjoy social chit-chat”), and imagination (e.g., “When I’m reading a story, I can easily imagine what the characters might look like”). Each item is rated on a 4-point Likert scale ranging from “definitely agree” to “definitely disagree.” Scoring is binary (0– 1): 24 positively worded items are scored 1 point if the respondent selects “definitely agree” or “slightly agree,” and 0 for “slightly disagree” or “definitely disagree.” The remaining 26 reverse-scored items are coded oppositely. Higher total scores indicate stronger autistic traits. In the present study, the AQ demonstrated high internal consistency, with a Cronbach’s alpha coefficient of 0.932 for the total scale. The sub-scales also showed acceptable reliability, with Cronbach’s α of 0.734 for social skills, 0.713 for attention switching, 0.754 for attention to detail, 0.749 for communication, and 0.704 for imagination.

### Research Results

All data were organized and analyzed using SPSS version 26.0.

### Descriptive Statistical Analysis

The average Autism-Spectrum Quotient (AQ) score among the college student participants was 21.21 (*SD* = 5.27), indicating a moderate level of autistic traits overall. A total of 90 individuals scored above 28, accounting for 9.93% of the total sample. Scores on the individual sub-scales ranged between 0 and 10. Detailed descriptive statistics are presented in Table 1.

**Table 1.** Descriptive statistical analysis.

| Item | Minimum | maximum | $M \pm SD$ |
| --- | --- | --- | --- |
| AQ | 7 | 38 | 21.21 ± 5.27 |
| **social skills** | 0 | 10 | 3.47 ± 2.31 |
| **attention switching** | 1 | 10 | 5.10 ± 1.54 |
| **attention to detail** | 0 | 10 | 5.43 ± 1.90 |
| **communication** | 0 | 9 | 3.65 ± 2.01 |
| **imagination** | 0 | 8 | 3.57 ± 1.69 |

### Analysis of Group Differences

An independent samples *t*-test revealed a significant main effect of gender on the total AQ score, with males scoring significantly higher than females. Regarding the five AQ subscales (Table 2), significant gender differences were observed in the dimensions of communication and imagination, where males scored notably higher than females (ps < 0.001). No significant gender differences were found in the remaining subscales.

**Table 2.** Analysis of differences between different genders.

| Item | Group | *M* ± *SD* | *t* | Cohen's *d* |
| --- | --- | --- | --- | --- |
| **AQ** | Male | 21.57 ± 5.13 | 2.52* | 0.170 |
|  | Female | 20.67 ± 5.44 |  |  |
| **Social skills** | Male | 3.45 ± 2.22 | -0.28 | 0.021 |
|  | Female | 3.50 ± 2.44 |  |  |
| **Attention switching** | Male | 5.04 ± 1.48 | -1.32 | 0.09 |
|  | Female | 5.18 ± 1.62 |  |  |
| **Attention to detail** | Male | 5.48 ± 1.90 | 0.86 | 0.063 |
|  | Female | 5.36 ± 1.89 |  |  |
| **Communication** | Male | 3.84 ± 1.99 | 3.51*** | 0.239 |
|  | Female | 3.36 ± 2.01 |  |  |
| **Imagination** | Male | 3.76 ± 1.65 | 4.34*** | 0.290 |
|  | Female | 3.27 ± 1.72 |  |  |

PS: In the above table, \*indicates $p < 0.05$ ; \*\*indicates $p < 0.01$ ; \*\*\*indicates that $p < 0.001$ , the same below.

### Study 2: The effect of Tai Chi intervention on autistic traits in college students—An analysis based on emotion regulation self-efficacy and aggressive behavior Participants

In accordance with previous research, individuals scoring above 28 on the Chinese version of the Autism-Spectrum Quotient (AQ) were identified as exhibiting high levels of autistic traits (Zhang et al., 2021). As shown in Fig 1, among the participants who have participated in Study 1, those who meet the requirements were selected. Based on this criterion and after obtaining informed consent, a total of 20 participants were recruited, including 15 males and 5 females, aged between 18 and 20 years (M = 18.65, SD = 0.59). Participants were randomly assigned to either the experimental group (Tai Chi intervention group, n = 10) or the control group (non-intervention group, n = 10). The participant recruitment period for this study was from October 14, 2024 to December 8, 2024. All procedures were approved by the Ethics Committee of Shandong University of Political Science and Law and were conducted in accordance with the Declaration of Helsinki. Participants voluntarily took part in the study and were required to read and confirm the informed consent form before completing the questionnaire and prior to the intervention phase. The study ensured confidentiality and allowed participants to withdraw at any time if they felt uncomfortable or for any other reason. Upon completion of the intervention, participants in the experimental group received a compensation of 100 RMB, while those in the control group received 10 RMB.

**Fig 1.**
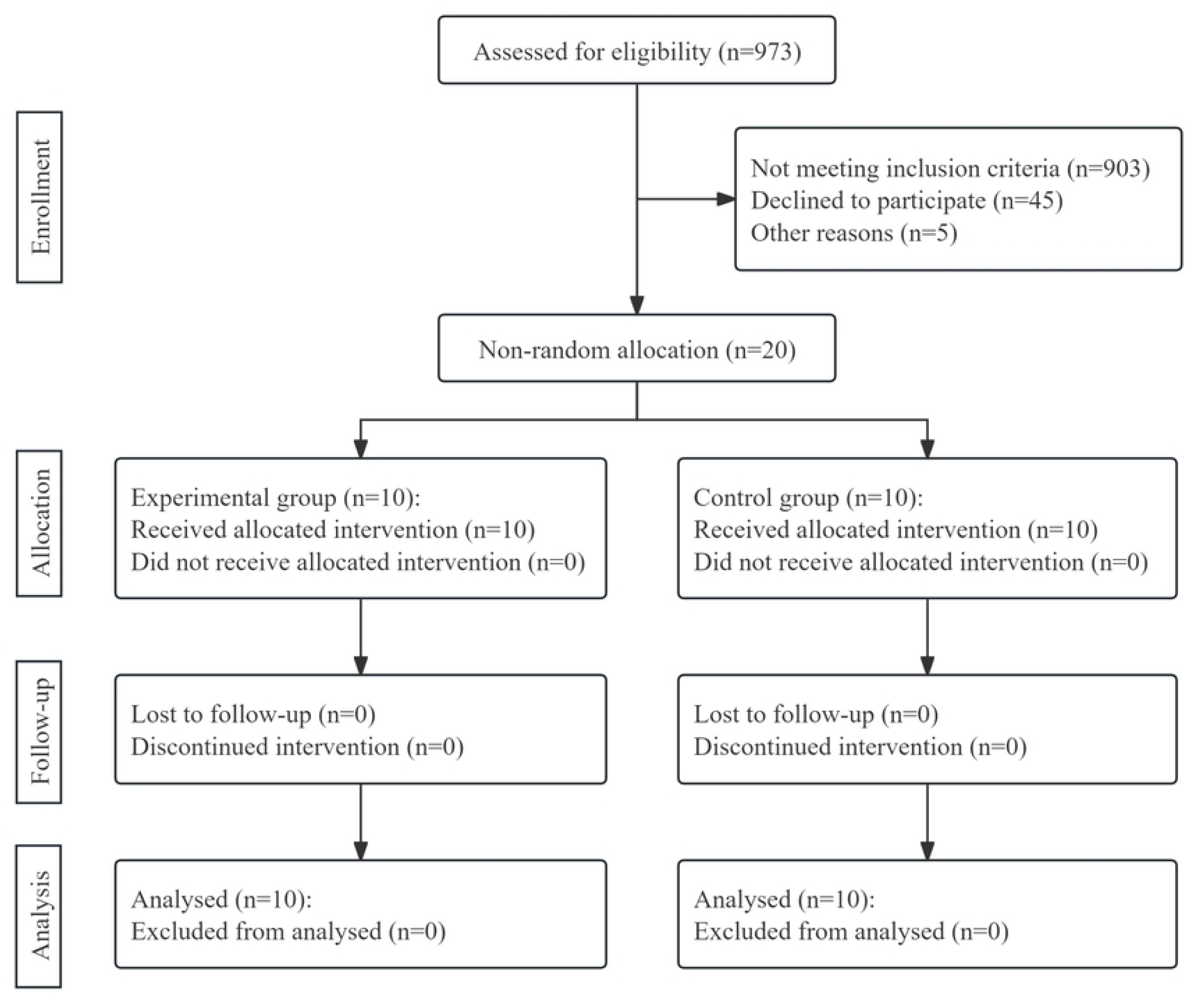
CONSORT flow diagram.

### Materials

(1) Autism-Spectrum Quotient (AQ)

The AQ was the same instrument used in Study 1.

(2) Regulatory Emotional Self-Efficacy Scale (RESE)

Originally developed by Bandura, Caprara (19) and adapted to Chinese by Zhang, Zhang (20), the RESE has demonstrated sound psychometric properties among Chinese college student populations. Higher total scores reflect stronger self-perceived efficacy in regulating emotional responses, indicating greater confidence and effectiveness in managing a range of emotional experiences. In the current study, the scale demonstrated good internal consistency, with a Cronbach’s α of 0.906.

(3) Reactive-Proactive Aggression Questionnaire (RPQ)

The Chinese version of the RPQ was revised by Zhang, Jia (21). This 20-item questionnaire measures two distinct dimensions of aggression: reactive aggression (e.g., “I have gotten angry when threatened by others”) and proactive aggression (e.g., “I have hit others to assert dominance”). These dimensions reflect different emotional and behavioral characteristics—reactive aggression is typically associated with anger, hostility, and impulsive responses to provocation, while proactive aggression refers to deliberate, goal-oriented aggressive acts without immediate provocation. Items assessing reactive aggression include items 1, 3, 5, 7, 8, 11, 13, 14, 18, and 20, while proactive aggression is measured by items 2, 4, 6, 9, 10, 12, 15, 16, 17, and 19. Responses are rated on a 6-point Likert scale ranging from 1 (Never) to 6 (Always). Higher scores on the reactive sub-scale indicate a greater tendency toward emotional and defensive responses when facing perceived threats, whereas higher proactive aggression scores reflect a greater likelihood of initiating aggression for instrumental purposes. In this study, the Cronbach’s α coefficients of the total scale, proactive aggression and reactive aggression were 0.975, 0.971 and 0.946 respectively.

### Experimental Procedure

A 2 (Group: Tai Chi group vs. control group) × 2 (Time: pre-intervention vs. post-intervention) mixed factorial design was adopted, with group as a between-subjects factor and time as a within-subjects factor. The dependent variables were participants’ self-reported scores on the respective scales.

The experimental procedure consisted of three phases: pre-intervention assessment, an 8-week intervention period, and post-intervention assessment. Prior to the intervention, participants provided demographic information (including age and gender) and completed all baseline psychological measures.

During the 8-week intervention phase, participants in the Tai Chi group practiced the standardized 24-form Chen-style Tai Chi, while the control group received no Tai Chi training and continued with their usual routines. The 24-form Chen-style Tai Chi is a simplified sequence developed after the founding of the People’s Republic of China. Originating in China, Tai Chi integrates elements from various traditional martial arts, ancient health exercises such as Daoyin and Qigong, as well as classical philosophy and traditional Chinese medicine. It is characterized by gentle, slow, and flowing movements that emphasize both internal and external coordination. The intervention was led by a certified martial arts instructor with 16 years of Tai Chi practice and a sixth-level national martial arts qualification. Participants in the intervention group were required to wear appropriate athletic clothing and shoes during each session.

All participants signed informed consent forms prior to the beginning of the intervention. The Tai Chi group participated in one 45-minute training session per week over 8 weeks. Each session comprised four parts: a 5-minute warm-up, 20 minutes of new movement instruction, 15 minutes of full-sequence practice, and a 5-minute cool-down. The warm-up focused on mobilizing the neck, wrists, and ankles to reduce injury risk. During the learning phase, the 24 Tai Chi movements were introduced and reviewed step-by-step. The full-sequence practice required participants to perform the complete routine twice, reinforcing the newly learned skills. The cool-down involved body tapping and breathing regulation, led by the instructor to promote relaxation and recovery.

Because none of the participants had prior experience with Tai Chi or other martial arts, a high-frame posture was used throughout the intervention to facilitate learning. The instructor alternated between mid-and high-frame styles when demonstrating movements, with mid-frame used for leading the group through the complete sequence.

After completing the Tai Chi intervention, all participants were required to complete the same set of questionnaires again. Upon finishing the entire experiment, they received compensation for their participation.

## Research Results

### Pre-Test Comparison Between Experimental and Control Groups

An independent samples t-test was conducted to examine the baseline equivalence of the Tai Chi group and the control group in terms of age, height, weight, and BMI. The results indicated no significant differences between the two groups on any of these demographic variables (see Table 3), suggesting there was no difference in the basic information of the participants before the intervention stage.

**Table 3.** Difference analysis of basic information.

| Item | Group | $M \pm SD$ | $t$ |
| --- | --- | --- | --- |
| Age | Tai Chi group | $18.60 \pm 0.67$ | -0.37 |
| | control group | $18.70 \pm 0.48$ | |
| Height/m | Tai Chi group | $1.75 \pm 0.05$ | 1.60 |
| | control group | $1.71 \pm 0.06$ | |
| Weight/kg | Tai Chi group | $69.60 \pm 5.25$ | 1.45 |
| | control group | $66.00 \pm 5.81$ | |
| BMI | Tai Chi group | $22.66 \pm 0.69$ | 0.57 |
| | control group | $22.47 \pm 0.81$ | |

### Post-intervention differences between Tai Chi and control groups

A 2 (Group: Tai Chi vs. Control) × 2 (Time: Pre-test vs. Post-test) repeated measures ANOVA was conducted to examine the intervention effects. As shown in Table 4, there was a significant main effect of time on the total AQ score, and the interaction between time and group was significant (*F* = 6.86, *p* < 0.05, *η^2^* = 0.276). Simple effect analysis revealed that the post-test AQ score in the experimental group was significantly lower than the pre-test score (*M* =-5.30, *SD* = 1.40, *p* < 0.01), indicating a notable reduction in autistic traits following the Tai Chi intervention.

**Table 4.** Analysis of the differences in AQ scores between different groups.

| Item | Group | $M \pm SD$ | $F$ | $\eta^2$ |
| --- | --- | --- | --- | --- |
| pre-AQ | Tai Chi | $31.30 \pm 1.57$ | 6.86* | 0.276 |
| | control | $31.00 \pm 1.56$ | | |
| post-AQ | Tai Chi | $26.00 \pm 4.35$ | | |
|  | control | 30.90 ± 2.13 |  |  |
| **pre-social skills** | Tai Chi | 6.10 ± 2.28 | 0.02 | 0.001 |
|  | control | 6.50 ± 1.78 |  |  |
| **post-social skills** | Tai Chi | 6.20 ± 2.74 |  |  |
|  | control | 6.80 ± 1.62 |  |  |
| **pre-attention switching** | Tai Chi | 7.40 ± 1.08 | 28.46*** | 0.613 |
|  | control | 6.50 ± 1.51 |  |  |
| **post-attention switching** | Tai Chi | 3.80 ± 1.48 |  |  |
|  | control | 6.60 ± 1.17 |  |  |
| **pre-attention to detail** | Tai Chi | 6.90 ± 1.66 | 6.34* | 0.260 |
|  | control | 6.10 ± 1.45 |  |  |
| **post-attention to detail** | Tai Chi | 4.00 ± 2.16 |  |  |
|  | control | 6.20 ± 2.04 |  |  |
| **pre-communication** | Tai Chi | 6.10 ± 1.45 | 0.06 | 0.004 |
|  | control | 5.90 ± 0.99 |  |  |
| **post-communication** | Tai Chi | 5.60 ± 2.37 |  |  |
|  | control | 5.10 ± 1.29 |  |  |
| **pre-imagination** | Tai Chi | 4.80 ± 1.81 | 1.73 | 0.088 |
|  | control | 6.00 ± 1.41 |  |  |
| **post-imagination** | Tai Chi | 6.40 ± 1.78 |  |  |
|  | control | 6.20 ± 1.23 |  |  |

Regarding AQ sub-dimensions, a significant time × group interaction was observed for the attention switching sub-scale (*F*=28.46, *p* < 0.001, *η^2^* = 0.61). Simple effect analysis indicated that the experimental group showed a significant decrease from pre-test to post-test (*M* =-3.60, *SD* = 0.49, *p* < 0.001), suggesting improved cognitive flexibility after the intervention. Similarly, for the attention to detail sub-scale, the interaction between time and group was significant (*F* = 6.34, *p* < 0.05, *η^2^* = 0.26). Simple effect analysis showed that the experimental group’s post-test scores were significantly lower than their pre-test scores (*M* =-2.90, *SD* = 0.84, *p* < 0.001), indicating reduced over-focus on details. In other dimensions, the interaction between time and group was not significant.

As presented in Table 5, in terms of the scores of the RESE, the interaction between time and group was significant (*F* = 8.87, *p* < 0.01, *η^2^* = 0.330). Simple effect analysis revealed that post-test scores in the experimental group were significantly higher than pre-test scores (*M* = 11.00, *SD* = 3.36, p < 0.01), indicating that participants’ perceived emotional self-efficacy significantly improved following the Tai Chi intervention.

**Table 5.**
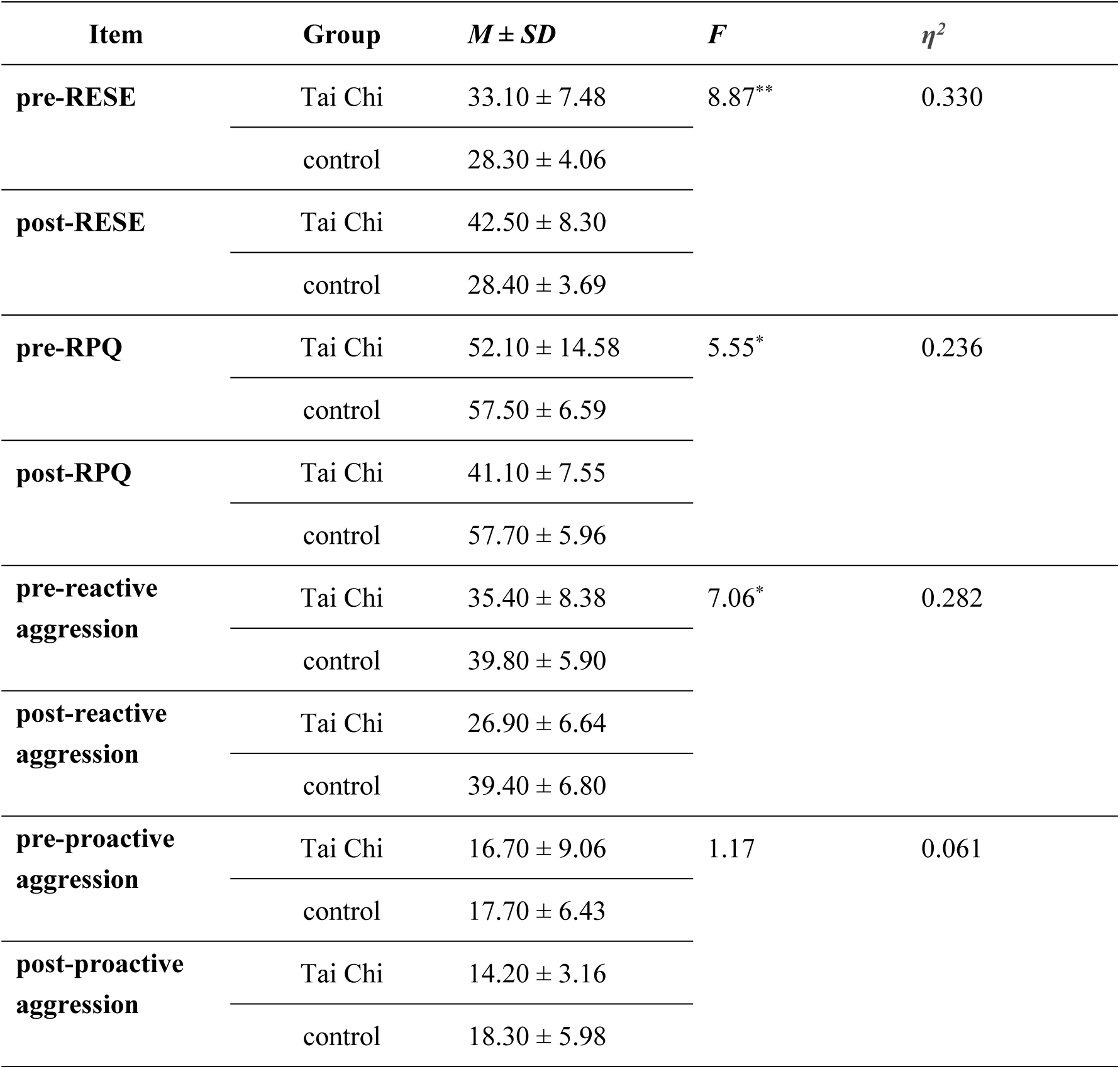
Analysis of the differences in RESE and RPQ scores between different groups.

For the score of RPQ, the interaction between time and group was also significant (*F* = 8.87, *p* < 0.01, *η^2^* = 0.330). Simple effect analysis showed that the experimental group exhibited significantly lower post-test scores compared to pre-test scores (*M* = 11.00, *SD* = 3.36, *p* < 0.01), suggesting a reduction in aggression-related behaviors due to the intervention. Specifically, for the reactive aggression sub-scale, the interaction between time and group was significant (*F* = 8.52, *p* < 0.01, *η2* = 0.32). Simple effect analysis indicated that the post-test scores of the experimental group were significantly lower than the pre-test scores (*M* =-8.50, *SD* = 2.16, *p* < 0.01), demonstrating that Tai Chi practice effectively reduced participants’ levels of reactive aggression.

For the score of RPQ, the interaction between time and group was also significant (*F* = 8.87, *p* < 0.01, *η^2^* = 0.330). Simple effect analysis showed that the experimental group exhibited significantly lower post-test scores compared to pre-test scores (*M* = 11.00, *SD* = 3.36, *p* < 0.01), suggesting a reduction in aggression-related behaviors due to the intervention. Specifically, for the reactive aggression sub-scale, the interaction between time and group was significant (*F* = 8.52, *p* < 0.01, *η^2^* = 0.32). Simple effect analysis indicated that the post-test scores of the experimental group were significantly lower than the pre-test scores (*M* =-8.50, *SD* = 2.16, *p* < 0.01), demonstrating that Tai Chi practice effectively reduced participants’ levels of reactive aggression.

## Discussion

### Analysis of the current status of autistic traits among college students

The first phase of this study examined the prevalence of autistic traits in the college student population and the differences based on gender. The results indicated that the average score of AQ among college students is 21.21, suggesting a moderate level of autistic traits. This finding supports the assertion by Ruzich, Allison (3) that autistic traits are continuously distributed in the general population, characterized by substantial individual variability, and can be conceptualized as a quantifiable dimension of personality. It further implies that autistic traits are not confined to clinical diagnoses of ASD but are also present, to varying degrees, in non-clinical populations. In particular, college students—who often experience complex social and academic demands—may exhibit more pronounced expressions of these traits.

Further analysis revealed that 90 individuals (9.93% of the sample) scored above the AQ threshold of 28, indicating a relatively elevated level of autistic traits. According to Baron-Cohen, Wheelwright (22), higher AQ scores reflect greater difficulties in areas such as social interaction, communication, and imagination. Although individuals with high AQ scores may not meet the diagnostic criteria for ASD, they are likely to encounter more psychological and social adaptation challenges in daily life. These findings underscore the importance of providing tailored psychological support services to this subgroup to enhance their overall well-being and functioning.

With regard to gender differences, the present study found that male participants scored significantly higher than female participants on the AQ, suggesting that males exhibit more pronounced autistic traits. This finding is consistent with previous research both in China and internationally. For example, a systematic review by Loomes, Hull (4) reported a notable gender disparity in clinical diagnoses of autism spectrum disorder (ASD), with male-to-female ratios ranging from approximately 3:1 to 4:1. This gender difference extends beyond clinical populations and is also evident in the distribution of autistic traits among the general population [3]. Such findings have prompted theoretical debates on the etiology and nature of ASD, including the extreme male brain hypothesis and the female protective effect [4]. On the one hand, biological mechanisms may contribute to this disparity. For instance, prenatal exposure to higher levels of androgens, such as testosterone, has been associated with increased systemizing tendencies and elevated autistic traits [23]. On the other hand, from the perspective of cognitive style theory, males are generally more inclined toward “systemizing”—the drive to analyze, construct, and understand rule-based systems—whereas females typically exhibit stronger empathizing skills, reflecting the capacity to understand and respond to others’ emotions [24]. These cognitive style differences may partially explain the higher AQ scores observed in males. Moreover, sociocultural factors should not be overlooked. Females may be more likely to mask or compensate for autistic traits due to social expectations, especially in non-clinical samples. As a result, conventional assessment tools such as the AQ may underestimate the level of autistic traits in females [25]. It is also noteworthy that the female phenotype of ASD may present more subtly, with fewer overt restricted interests and greater ability to camouflage symptoms, making ASD in females less likely to be recognized or diagnosed [4].

Further analysis of the five sub-scales of the AQ revealed that male participants scored significantly higher than females on the dimensions of communication and imagination. This suggests that males exhibit more pronounced characteristics associated with core features of the autism spectrum, specifically impairments in verbal communication and deficits in imaginative capacity. Previous research has shown that individuals on the autism spectrum often demonstrate marked challenges in communication, such as poor pragmatic language skills, difficulty interpreting non-literal expressions, reduced eye contact, and limited empathetic interaction [26]. Deficits in imagination are commonly reflected in reduced associative thinking, limited capacity for symbolic play, and a tendency toward repetitive behaviors [27]. The elevated scores in these areas among male college students may indicate greater difficulties in social adaptation, particularly in environments requiring effective teamwork, emotional expression, and creativity.

Overall, the findings of this study underscore two key points. First, autistic traits are relatively common among university students; while most individuals fall within a moderate range, nearly 10% exhibit high levels of such traits, potentially placing them at greater risk for psychological maladjustment. Second, gender plays a significant role in the expression of autistic traits, with notable differences observed in the communication and imagination dimensions. These results provide a theoretical foundation for the development of targeted mental health services in higher education. In particular, psychological assessments, individual identification procedures, and intervention strategies should account for both inter-individual trait differences and gender-specific expressions of autistic characteristics.

### The impact of Tai Chi training on autistic traits in college students

The results demonstrated that an 8-week Tai Chi intervention led to a significant reduction in autistic traits among university students, with notable improvements in the subdomains of attention switching and attention to detail. Moreover, participants who underwent the intervention exhibited a marked increase in emotional regulation self-efficacy and a significant decrease in aggressive behaviors, particularly in reactive aggression. These findings suggest that Tai Chi training exerts a beneficial effect on enhancing social functioning, cognitive flexibility, and emotional regulation. Importantly, the intervention appears to offer meaningful mental health benefits for university students exhibiting moderate to high levels of autistic traits.

Firstly, the significant reduction in total AQ scores suggests that Tai Chi training contributes to a comprehensive improvement in individuals’ autistic traits. Autistic traits represent a multidimensional construct, encompassing domains such as social skills, attention switching, attention to detail, communication abilities, and imagination [3]. These domains collectively influence an individual’s psychological functioning in social interaction, emotional regulation, and information processing. As a mind–body integrative practice, Tai Chi emphasizes breath control, bodily awareness, and intentional focus, which may enhance perceptual sensitivity and self-awareness [28]. This mechanism may help individuals break out of entrenched patterns of cognitive rigidity and social withdrawal commonly associated with autistic traits, thereby contributing to an overall reduction in AQ scores. Further analysis revealed that participants in the Tai Chi group exhibited significant decreases in the subscales of attention switching and attention to detail. These findings indicate that Tai Chi may foster improvements in cognitive flexibility and holistic information processing. Individuals with higher autistic traits often display cognitive inflexibility and a local processing bias—characterized by excessive focus on details and difficulty in shifting attention between tasks [29]. Tai Chi training involves continuous modulation of attention during movement transitions, rhythm perception, and intentional guidance, requiring practitioners to adapt attentively at the somatic level [30]. This repetitive training process subtly cultivates the ability to integrate information in dynamic contexts, which may counteract rigid and stereotyped cognitive styles, as reflected in the observed subscale score reductions [30]. Moreover, Tai Chi emphasizes the unity of body and mind, encouraging practitioners to engage in the coordination of intention-driven movement(*心随意动、意动形随*),wherein mental focus guides physical actions. This process fosters the development of top-down attentional control—that is, attention guided by internal goals rather than external stimuli [31]. Enhanced attentional regulation through this mechanism may alleviate difficulties in attention switching and reduce overreliance on detailed information [32].

The enhancement of regulatory emotional self-efficacy represents another significant finding of this study, with important implications. Regulatory emotional self-efficacy refers to an individual’s confidence in their ability to manage and regulate negative emotional states, and improvements in this capacity have been shown to directly contribute to the reduction of anxiety, depression, and aggressive behavior [15, 33]. Tai Chi, characterized by its slow, harmonious movements and rhythmic breathing patterns, offers practitioners a setting conducive to deep relaxation and self-regulation [34]. This embodied experience promotes emotional introspection, enabling individuals to better recognize and modulate their emotional responses. Moreover, Tai Chi practice has been found to improve autonomic balance by enhancing the coordination between the sympathetic and parasympathetic nervous systems, thereby supporting the physiological basis of emotional regulation [35]. The positive somatic feedback generated during practice, such as slower respiration, muscle relaxation, and inner calmness, that fosters a heightened sense of self-regulatory control. As these internal experiences accumulate over time, they contribute to the progressive strengthening of one’s belief in their emotional regulatory abilities.

Finally, this study also revealed that participants in the intervention group exhibited significantly lower scores on the RPQ and the reactive aggression subscale. This suggests that Tai Chi practice may effectively reduce impulsive aggressive behaviors triggered by negative emotional states. Reactive aggression typically arises from emotional dysregulation in response to provocation and is closely associated with an individual’s emotional regulation capacity [2]. In China, university students often face high levels of academic competition, complex interpersonal dynamics, and transitional life challenges, all of which contribute to emotional instability [36]. Within such contexts, reactive aggression may emerge as a maladaptive coping strategy to mitigate psychological distress [33]. For college students, these aggressive responses are frequently driven by emotional arousal, particularly anger and frustration [37]. Given that university students are in a developmental stage characterized by ongoing identity formation and maturing emotional control, their capacity for emotional regulation may be underdeveloped, increasing vulnerability to emotionally driven aggression during interpersonal conflict or environmental stress. Moreover, under the influence of collectivist cultural norms that emphasize social harmony and group cohesion, Chinese students may be more inclined to suppress negative emotions in conflict situations [38]. However, such emotional suppression, if not effectively managed, can lead to abrupt outbursts of reactive aggression [39]. In this study, the observed improvements in regulatory emotional self-efficacy suggest that individuals became better equipped to regulate their emotional responses in challenging situations, thereby reducing the likelihood of impulsive aggressive reactions. In contrast, no significant changes were observed in proactive aggression, which is consistent with theoretical expectations. Proactive aggression is primarily a goal-oriented and cognitively mediated behavior—often motivated by dominance or instrumental gain—and is less dependent on immediate emotional states [12]. Therefore, short-term interventions are more likely to impact emotion-driven aggression rather than alter more stable behavioral patterns such as proactive aggression.

In summary, Tai Chi training appears to facilitate the integration and enhancement of individual psychological functioning through a mind–body synchronization mechanism across multiple dimensions. From a neurophysiological perspective, Tai Chi practice may enhance cognitive control and emotional inhibition by activating prefrontal cortical regions [40]; Additionally, the deliberate regulation of breathing and attention during training contributes to increased neural plasticity and improved autonomic nervous system regulation [35]; On a psychosocial level, group-based Tai Chi sessions provide a calm and inclusive social environment, offering individuals with high autistic traits opportunities for low-pressure social interaction. This may help reduce social anxiety and strengthen their sense of belonging [41]. Overall, the findings of this study suggest that Tai Chi should not merely be viewed as a traditional physical exercise, but rather as an integrative mind–body intervention that combines movement, breath control, and focused intention. Such a holistic approach demonstrates multiple psychological benefits, including improvements in autistic traits, enhanced emotional regulation efficacy, and reductions in aggressive behaviors among university students. Future research is warranted to further investigate the neural mechanisms underlying these effects and to assess the long-term impact and generalizability of Tai Chi across larger and more diverse populations.

## Conclusion and Limitations

This study explored the current status of autistic traits among university students and examined the effects of Tai Chi intervention through a combined approach of questionnaire-based assessment and experimental manipulation. It systematically investigated gender differences in autistic trait distribution and the positive effects of Tai Chi training on reducing autistic traits, enhancing emotional self-efficacy, and decreasing aggressive behaviors. The findings revealed that overall autistic trait levels among college students were within a moderate range, with males scoring significantly higher than females in total AQ scores as well as in the domains of communication and imagination. After participating in Tai Chi training, participants showed significant reductions in total AQ scores, particularly in attention switching and attention to detail subscales. Additionally, emotional regulation self-efficacy was significantly improved, and reactive aggression scores were markedly reduced, suggesting that Tai Chi has substantial value as a psychological intervention to promote mental health in university populations.

Despite the encouraging findings, several limitations should be acknowledged. First, the sample was relatively homogeneous, primarily drawn from universities in a specific geographic region, which may limit the generalizability of the results. Future studies should aim to include a more diverse and representative sample to enhance the external validity of the conclusions. Second, the intervention period was relatively short, and the study lacked long-term follow-up data, making it difficult to assess the durability of the intervention effects. Longitudinal designs are recommended for future research. Third, the underlying mechanisms of the intervention were not explored at the neural or physiological levels. Future studies may consider employing multimodal approaches, such as incorporating electroencephalography (EEG) and physiological indicators, to further elucidate the neurobiological mechanisms through which Tai Chi alleviates autistic traits.

## Declarations

### Ethics approval and consent to participate

All procedures were approved by the Ethics Committee of Shandong University of Political Science and Law and were conducted in accordance with the Declaration of Helsinki. Participants voluntarily took part in the study and were required to read and confirm the informed consent form before completing the questionnaire and prior to the intervention phase. The study ensured confidentiality and allowed participants to withdraw at any time if they felt uncomfortable or for any other reason.

## Consent for publication

Not applicable

## Availability of data and materials

The datasets generated and analyzed during the current study are not publicly available due to privacy issues. However, data will be available upon formal request to the corresponding author.

## Authors’ contributions

Yuan Zhao: Writing – review & editing, Writing – original draft, Visualization, Software, Methodology, Investigation, Formal analysis, Conceptualization. Yue Wang: Writing – review & editing, Validation, Supervision, Resources, Methodology, Data curation, Conceptualization. Weijiao Zhong: Validation, Supervision, Resources, Methodology. Qiqi Xing: Validation, Supervision, Methodology, Conceptualization.

## Acknowledgments

The authors would like to thank Zhuohao Wang, Yiming Wang, Jianming Guo, Zhenghao Yan, Guoshan Huang, Fengqi Yu, Zizhen Xu, Quanxin Wei and Zhemingjun Liu from Shandong University of Political Science and Law for their contributions to data collection and other research assistance. Best wishes to these valued colleagues - your support is deeply appreciated by the research team.

## References

1. Bloch C, Burghof L, Lehnhardt F-G, Vogeley K, Falter-Wagner C. Alexithymia traits outweigh autism traits in the explanation of depression in adults with autism. Scientific reports. 2021;11(1):2258.

2. Tan C, Song H, Ma S, Liu X, Zhao Y. Autistic Traits and Aggressive Behavior in Chinese College Students: A Serial Mediation Model and the Gender Difference. Psychology Research and Behavior Management. 2024:1385–97. doi: 10.2147/PRBM.S451028.

3. Ruzich E, Allison C, Smith P, Watson P, Auyeung B, Ring H, et al. Measuring autistic traits in the general population: a systematic review of the Autism-Spectrum Quotient (AQ) in a nonclinical population sample of 6,900 typical adult males and females. Molecular autism. 2015;6:1–12.

4. Loomes R, Hull L, Mandy WPL. What is the male-to-female ratio in autism spectrum disorder? A systematic review and meta-analysis. Journal of the American Academy of Child & Adolescent Psychiatry. 2017;56(6):466–74.

5. Lord C, Jones RM. Annual Research Review: Re-thinking the classification of autism spectrum disorders. Journal of Child Psychology and Psychiatry. 2012;53(5):490–509.

6. Ming X, Brimacombe M, Chaaban J, Zimmerman-Bier B, Wagner GC. Autism spectrum disorders: concurrent clinical disorders. Journal of child neurology. 2008;23(1):6–13.

7. Guo J, Shen Y, Li B, Wang F, Jiang Y, Lin Y, et al. Does Tai Chi Chuan improve psychological well-being and quality of life in patients with breast cancer? Protocol for a systematic review of randomized controlled trials: A Protocol for Systematic Review and Meta-Analysis. Medicine. 2020;99(16):e19681.

8. Chen J, Li Y, Wu Y, Su X. The influence of Tai Chi exercise on depressive mood and serum inflammatory factors of female college students. Chinese Journal of School Health. 2019;40(07):1065–8.

9. Jia X, Jiang C, Tao J, Li Y, Zhou Y, Chen L-d. Effects of core strength training combined with Tai Chi Chuan for the musculoskeletal system and cardiopulmonary function in older adults: A study protocol for a randomized controlled trial. Medicine. 2018;97(35):e12024.

10. Zhang S, Zou L, Chen L-Z, Yao Y, Loprinzi PD, Siu PM, et al. The effect of Tai Chi Chuan on negative emotions in non-clinical populations: a meta-analysis and systematic review. International Journal of Environmental Research and Public Health. 2019;16(17):3033.

11. Anderson CA, Bushman BJ. Human aggression. Annual review of psychology. 2002;53(1):27–51. doi: 10.1146/annurev.psych.53.100901.135231.

12. Raine A, Dodge K, Loeber R, Gatzke-Kopp L, Lynam D, Reynolds C, et al. The reactive–proactive aggression questionnaire: Differential correlates of reactive and proactive aggression in adolescent boys. Aggressive Behavior: Official Journal of the International Society for Research on Aggression. 2006;32(2):159–71.

13. Heynen E, Van der Helm P, Stams GJ, Roest J. Measuring aggression in German youth prison—A validation of the German Reactive-Proactive Aggression Questionnaire (RPQ) in a sample of incarcerated juvenile offenders. International journal of offender therapy and comparative criminology. 2022;66(13-14):1475–86.

14. Farmer CA, Aman MG. Aggressive behavior in a sample of children with autism spectrum disorders. Research in Autism Spectrum Disorders. 2011;5(1):317–23.

15. Naeem M, Weng Q, Ali A, Hameed Z. Linking family incivility to workplace incivility: Mediating role of negative emotions and moderating role of self-efficacy for emotional regulation. Asian Journal of Social Psychology. 2020;23(1):69–81.

16. Caprara GV, Di Giunta L, Eisenberg N, Gerbino M, Pastorelli C, Tramontano C. Assessing regulatory emotional self-efficacy in three countries. Psychological assessment. 2008;20(3):227.

17. Mazefsky CA, Herrington J, Siegel M, Scarpa A, Maddox BB, Scahill L, et al. The role of emotion regulation in autism spectrum disorder. Journal of the American Academy of Child & Adolescent Psychiatry. 2013;52(7):679–88.

18. Samson AC, Hardan AY, Podell RW, Phillips JM, Gross JJ. Emotion regulation in children and adolescents with autism spectrum disorder. Autism Research. 2015;8(1):9–18.

19. Bandura A, Caprara GV, Barbaranelli C, Gerbino M, Pastorelli C. Role of affective self-regulatory efficacy in diverse spheres of psychosocial functioning. Child development. 2003;74(3):769–82.

20. Zhang P, Zhang M, Lu J-M. An analysis of the results of the regulatory emotional self-efficacy scale in Chinese University students. Chinese Journal of Clinical Psychology. 2010. doi: 10.16128/j.cnki.1005-3611.2010.05.017.

21. Zhang WL, Jia S, Chen G, Zhang W. Reliability and validity of reactive-proactive aggression questionnaire in college students. Chinese Journal of Clinical Psychology. 2014;22(2):260–3. doi: 10.16128/j.cnki.1005-3611.2014.02.022.

22. Baron-Cohen S, Wheelwright S, Skinner R, Martin J, Clubley E. The autism-spectrum quotient (AQ): Evidence from asperger syndrome/high-functioning autism, malesand females, scientists and mathematicians. Journal of autism and developmental disorders. 2001;31:5–17. doi: 10.1023/A:1005653411471.

23. Baron-Cohen S. The extreme male brain theory of autism. Trends in cognitive sciences. 2002;6(6):248–54.

24. Lai M-C, Lombardo MV, Chakrabarti B, Ecker C, Sadek SA, Wheelwright SJ, et al. Individual differences in brain structure underpin empathizing–systemizing cognitive styles in male adults. Neuroimage. 2012;61(4):1347–54.

25. Lai M-C, Lombardo MV, Auyeung B, Chakrabarti B, Baron-Cohen S. Sex/gender differences and autism: setting the scene for future research. Journal of the American Academy of Child & Adolescent Psychiatry. 2015;54(1):11–24.

26. Barokova MD, La Valle C, Hassan S, Lee C, Xu M, McKechnie R, et al. Eliciting language samples for analysis (ELSA): A new protocol for assessing expressive language and communication in autism. Autism research. 2021;14(1):112–26.

27. Vyshedskiy A, editor Imagination in autism: a chance to improve early language therapy. Healthcare; 2021: MDPI.

28. Zou L, Loprinzi PD, Yu JJ, Yang L, Li C, Yeung AS, et al. Superior effects of modified chen-style tai chi versus 24-style tai chi on cognitive function, fitness, and balance performance in adults over 55. Brain sciences. 2019;9(5):102.

29. Happé F, Frith U. The weak coherence account: detail-focused cognitive style in autism spectrum disorders. Journal of autism and developmental disorders. 2006;36:5–25. doi: 10.1007/s10803-005-0039-0.

30. Hua H, Zhu D, Wang Y. Comparative study on the joint biomechanics of different skill level practitioners in Chen-Style Tai Chi punching. International Journal of Environmental Research and Public Health. 2022;19(10):5915.

31. Li Y, Chen X, Zhang Q, Xu W, Li J, Ji F, et al. Effects of working memory span training on top-down attentional asymmetry at both neural and behavioral levels. Cerebral Cortex. 2023;33(10):5937–46.

32. Tang Y-Y, Hölzel BK, Posner MI. The neuroscience of mindfulness meditation. Nature reviews neuroscience. 2015;16(4):213–25.

33. Liu H, Dou K, Yu C, Nie Y, Zheng X. The relationship between peer attachment and aggressive behavior among Chinese adolescents: the mediating effect of regulatory emotional self-efficacy. International journal of environmental research and public health. 2021;18(13):7123.

34. Dong Y, Pang D, Xiang J, Chao G, Kuang X. Exploring the benefits of traditional Chinese exercises (Tai Chi and Qigong) on the anxiety and depression of older adults: A systematic review and meta-analysis. Medicine. 2025;104(12):e41908.

35. Larkey L, James D, Vizcaino M, Kim SW. Effects of Tai Chi and Qigong on Heart Rate Variability: A Systematic Review and Meta-Analysis. Heart and Mind. 2024;8(4):310–24.

36. Guo J, Tang X, Marsh HW, Parker P, Basarkod G, Sahdra B, et al. The roles of social–emotional skills in students’ academic and life success: A multi-informant and multicohort perspective. Journal of personality and social psychology. 2023;124(5):1079.

37. Chen C, Chen X, Hou S, Lin S, Natthamon K. ANALYSIS ON THE CAUSES OF ANXIETY SENSITIVITY OF POST MILLENNIUM COLLEGE STUDENTS’AGGRESSIVE BEHAVIOR IN THE CONTEXT OF INTEGRATED MEDIA. International Journal of Neuropsychopharmacology. 2022;25(Supplement_1):A7-A.

38. Liw L, Ciftci A, Kim T. Cultural values, shame and guilt, and expressive suppression as predictors of depression. International Journal of Intercultural Relations. 2022;89:90–9.

39. Gutiérrez-Cobo MJ, Megías-Robles A, Gómez-Leal R, Cabello R, Fernández-Berrocal P. Emotion regulation strategies and aggression in youngsters: The mediating role of negative affect. Heliyon. 2023;9(3).

40. Yu AP, Tam BT, Lai CW, Yu DS, Woo J, Chung K-F, et al. Revealing the neural mechanisms underlying the beneficial effects of Tai Chi: a neuroimaging perspective. The American journal of Chinese medicine. 2018;46(02):231–59.

41. Yang G, Li X, Peel N, Klupp N, Liu J-P, Bensoussan A, et al. Perceptions of participants on trial participation and adherence to Tai Chi: a qualitative study. Patient preference and adherence. 2022:2695–707.

